# Evaluation of SARS-CoV-2 antibody response between paired fingerprick (hemaPEN®) and venepuncture collected samples in children and adults

**DOI:** 10.1101/2024.07.24.24310954

**Authors:** Nadia Mazarakis, Zheng Quan Toh, Jill Nguyen, Rachel A. Higgins, James Rudge, Belinda Whittle, Nicholas J. Woudberg, Justin Devine, Andrew Gooley, Florian Lapierre, Nigel W. Crawford, Shidan Tosif, Paul V Licciardi

## Abstract

Serological surveillance of severe acute respiratory syndrome coronavirus 2 (SARS-CoV-2) antibodies is important to monitor population COVID-19 immunity. Dried blood spots (DBS) are a valuable method for serosurveys, particularly in remote settings and in children. We compared the measurement of SARS-CoV-2 spike-specific IgG in paired blood samples collected by standard venepuncture (serum) and the hemaPEN® microsampling DBS device from children and adults using an established in-house ELISA. A total of 83 participants (10 months – 65 years of age) with paired serum and hemapen samples were included in the analysis; N=41 adults (36 COVID-positive and 5 COVID-negative) and N=42 children (37 COVID positive and 5 COVID-negative). Moderate-to strong-correlations between paired hemaPEN DBS eluates and serum SARS-CoV-2 IgG antibodies for RBD (r=0.9472, P<.0001) and S1 proteins (r=0.6892, P<.0001) were found. Similar results were observed in both adult and paediatric populations. SARS-CoV-2 spike-specific IgG in hemaPEN DBS samples remained stable for at least 35 weeks at room temperature. HemaPEN samples showed high specificity and sensitivity (100% and 89.89%, respectively) compared with serum. The use of the microsampling hemaPEN device for DBS sample collection is a feasible approach for assessing SARS-CoV-2 antibodies for serosurveillance studies, particularly in remote settings and in children.

## Introduction

Serological surveillance of severe acute respiratory syndrome coronavirus 2 (SARS-CoV-2) can provide estimates of population exposure and immunity, an important component to help inform the public health response. The emergence of new SARS-CoV-2 variants that escape immunity from earlier variants and waning immunity justifies the need for surveillance across the life course, with Australia having high seroprevalence following the Omicron wave in late 2021, early 2022 [1]. Continued serosurveillance is particularly important in countries with low vaccine coverage and in young infants < 2 years, who may not have been exposed to the SARS-CoV-2 virus.

Serology testing typically involves serum/plasma samples collected by venepuncture. However, this method requires access to trained staff, blood processing equipment, -20°C freezer infrastructure, and transportation logistics to laboratory facilities, all of which can be costly and logistically challenging, especially in lower to middle income countries (LMIC) and remote settings. Fingerprick tests on dried blood spot (DBS) cards is a logistically feasible and less invasive approach than venepuncture. The DBS approach has been used to measure serological response in a number of infectious diseases and vaccines including measles [2], mumps, and rubella [3, 4]. Furthermore, we and several other studies have demonstrated the measurement of SARS-CoV-2 antibodies in DBS collected on Gutherie cards [5-10]. However, there are some limitations to DBS samples collected on Guthrie cards such as sample volume variability, requirements to wait for cards to dry and appropriate storage to prevent contamination risk.

The hemaPEN® is a novel microsampling device for the collection of DBS samples offers some advantages over conventional DBS. The hemaPEN device uses capillary mechanism to collect four volumetrically fixed DBS samples, in a self-contained and easily storable way which reduces the risk of contamination.

This study aimed to evaluate the use of the hemaPEN microsampling device for SARS-CoV-2 antibodies against paired venepuncture serum samples in adult and paediatric populations following SARS-CoV-2 infection and/or COVID-19 vaccination, using an established in-house ELISA. A secondary aim was to determine whether a Synexa assay platform can detect Spike-specific IgG and neutralising antibodies using DBS from the hemaPEN microsampling device and how well the Synexa assay correlates with the in-house ELISA.

## Methods

### Study cohort

Participants were recruited as part of a household cohort study if they presented with a SARS-CoV-2 positive test (nasal/throat swab PCR-positive) at the Royal Children’s Hospital, Australia [11]. We also recruited participants who were SARS-CoV-2 negative and/or COVID-19 vaccinated only from the household cohort and a Health Care Worker vaccine cohort [12]. In both studies, data on COVID-19 PCR testing and vaccination records were collected. All COVID-19 among participants in our study were mild and did not require hospitalisation.

Written informed consent and assent were obtained from adults/parents and children, respectively. Participants provided a fingerprick hemaPEN DBS sample as well as a venepuncture blood sample. This study was approved by the Royal Children’s Hospital Melbourne Human Research Ethics Committee (HREC): HREC/63666 and HREC 74705/RCHM-2019.

### Sample collection and storage

For the fingerprick sample collection, an alcohol swab was used to sanitise the finger, followed by a lancet to create a drop of blood on the finger. The blood was collected by a hemaPEN device that consists of four capillary tubes that transfer the blood onto four DBS discs, equivalent to 2.74μL of blood/disc. HemaPEN collected samples were stored at room temperature at MCRI until analysis. A standard venepuncture procedure was used to collect blood samples. Blood sample was collected using SST gel tubes (Sarstedt, Nümbrecht Germany) and was processed within 2 hours of collection to obtain serum. The serum samples were stored at -80°C.

### Measurement of SARS-CoV-2 RBD- or S1-IgG using in-house ELISA

Two DBS discs collected from the hemaPEN device were diluted in a total volume of 274μL 10% skim milk-PBS-0.1% Tween (1:50). Samples were eluted overnight with shaking at 500 rpm at room temperature [8]. An in-house SARS-CoV-2 ELISA was used for this study, which was previously compared with commercial assays including DiaSorin LIAISON® SARS-CoV-2 IgG Test and Wantai SARS-CoV-2 ELISA kit as previously reported [8, 11]. Briefly, high-binding 96-well plates were coated with 2 μg/mL of receptor-binding domain (RBD), or S1 protein of the ancestral strain (Sino Biological, Beijing, China) diluted in phosphate-buffered saline (PBS). Plates were blocked with 10% skim milk-PBS-0.1%Tween, prior to the addition of paired serum (1:50) and hemaPEN DBS eluates (1:50) samples. A goat anti-human IgG (1:10,000) horseradish peroxidase–conjugated secondary antibody was used, and the plates were developed using 3.3⍰, 5.5⍰-tetramethylbenzidine substrate solution. Samples were first screened with the SARS-CoV-2 RBD antigen, and seropositive samples were confirmed and titrated with the S1 antigen. For RBD, results were reported as optical density (OD450nm), with results greater than 0.5 OD units considered seropositive based on pre-pandemic controls [8]. For S1, results were converted to binding antibody units (BAU/ml) based on a World Health Organization SARS-CoV-2 pooled serum standard (National Institute of Biological Standards and Controls, UK). The cut-off for seropositivity was 8.82 BAU/mL. The cut-offs values for RBD and S1 were based on our previously optimised methodology that demonstrated a strong correlation between paired DBS and serum samples [8].

### Measurement of SARS-CoV-2 antibodies using Synexa assay

A total of N=150 DBS discs were sent to Synexa (South Africa) from N=83 participants (up to two hemaPEN DBS discs per participant) for measurement of SARS-CoV-2 antibodies. Analysis of samples at Synexa was undertaken 32 weeks after collection and were stored at Synexa at -20°C prior to analysis. Control samples (N=7 negative SARS-CoV-2 and N=3 convalescent hemaPEN samples) were provided by Synexa in addition to the DBS samples.

Samples and controls were eluted according to Synexa’s DBS sample processing protocol as detailed in Maritz et al. 2022 [6]. Briefly, the DBS samples were eluted into LowCross BufferR® (Candor Bioscience; Germany) at 350 ± 50 RPM overnight at 2–8⍰C and following elution, stored at-70⍰C before analysis. The Synexa assay measures IgG specific for the SARS-CoV-2 Spike protein and neutralising antibodies (NAb) using a surrogate neutralising competitive assay. Assay wells were coated with the wildtype S1 spike protein. The IgG-specific assay used an anti-human IgG secondary antibody for detection. The neutralising assay detected antibodies with HRP-conjugated ACE-2. These assays are semi-quantitative, with an assay-cut-point (ACP) calculated based off the mean signal of the negative controls, as previously described [6]. Binding IgG antibodies were considered positive above the ACP cut-off, and for NAb, signals below the ACP cut-off were deemed positive.

### Statistical analysis

The RBD and S1 IgG concentrations of paired hemaPEN DBS eluates and serum samples were compared using a non-parametric Wilcoxon matched pairs signed ranked test. For correlation analyses, a Pearson’s correlation was used for RBD and S1 IgG concentrations, for in-house ELISA and Synexa assay. The level of agreement between the hemaPEN DBS eluates and serum samples was assessed by Bland-Altman analysis, and the sensitivity and specificity were reported. Statistical analyses were performed using GraphPad Prism 9.1.1 (GraphPad Software Inc., San Diego, U.S.), with a two-sided p<0.05 considered statistically significant.

## Results

### Characteristics of participants

A total of N=83 participants (10 months – 65 years of age) were recruited (Table 1) between July and December 2021. Participants were defined as positive for SARS-CoV-2 antibodies either by infection defined as RT-PCR positive, history of COVID-19 vaccination, or a combination of infection and vaccinated. Additionally, participants that were defined as negative for SARS-CoV-2 antibodies were confirmed by RT-PCR and grouped as uninfected/unvaccinated.

**Table 1:** Participant’s Characteristics.

| Table 1: Participant's Characteristics |  |  |
| --- | --- | --- |
|  | Adult (N=41) | Children (N=42) |
| Age in years, median (IQR) | 38 (26-65) | 5 (0-18) |
| Infected, N (%) | 13 (31.7%) | 36 (85.7%) |
| Time since last documented infection (weeks), median (IQR) | 52 (51.9-52.1) | 5.1 (4.6-7.3) |
| Vaccinated, N (%) | 16 (39%) | 1 (2.4%) |
| Time since last vaccine dose (weeks), median (IQR) | 6.3 (4.0-13.5) | 8.1 |
| Infected and Vaccinated, N (%) | 7 (17.1%) | - |
| Negative, N (%) | 5 (12.2%) | 5 (11.9%) |

### Comparison of SARS-CoV-2 RBD and S1 antibody responses between hemaPEN DBS eluates and serum samples

Between paired serum and hemaPEN DBS eluates, the proportion of seropositive samples were not significantly different for RBD and S1 (Fig. 2 A and D, respectively). The hemaPEN DBS eluates had significantly lower IgG concentrations specific for RBD and S1 (P< .0001) compared to serum (Fig.2. B and E, respectively). Moderate to strong correlations were observed for both SARS-CoV-2 RBD (r=0.9472, P <.0001) and S1-specific IgG S1 (r=0.6892, P <.0001) between paired hemaPEN eluted DBS and serum (Fig. 2 C and F, respectively).

**Figure 1.**
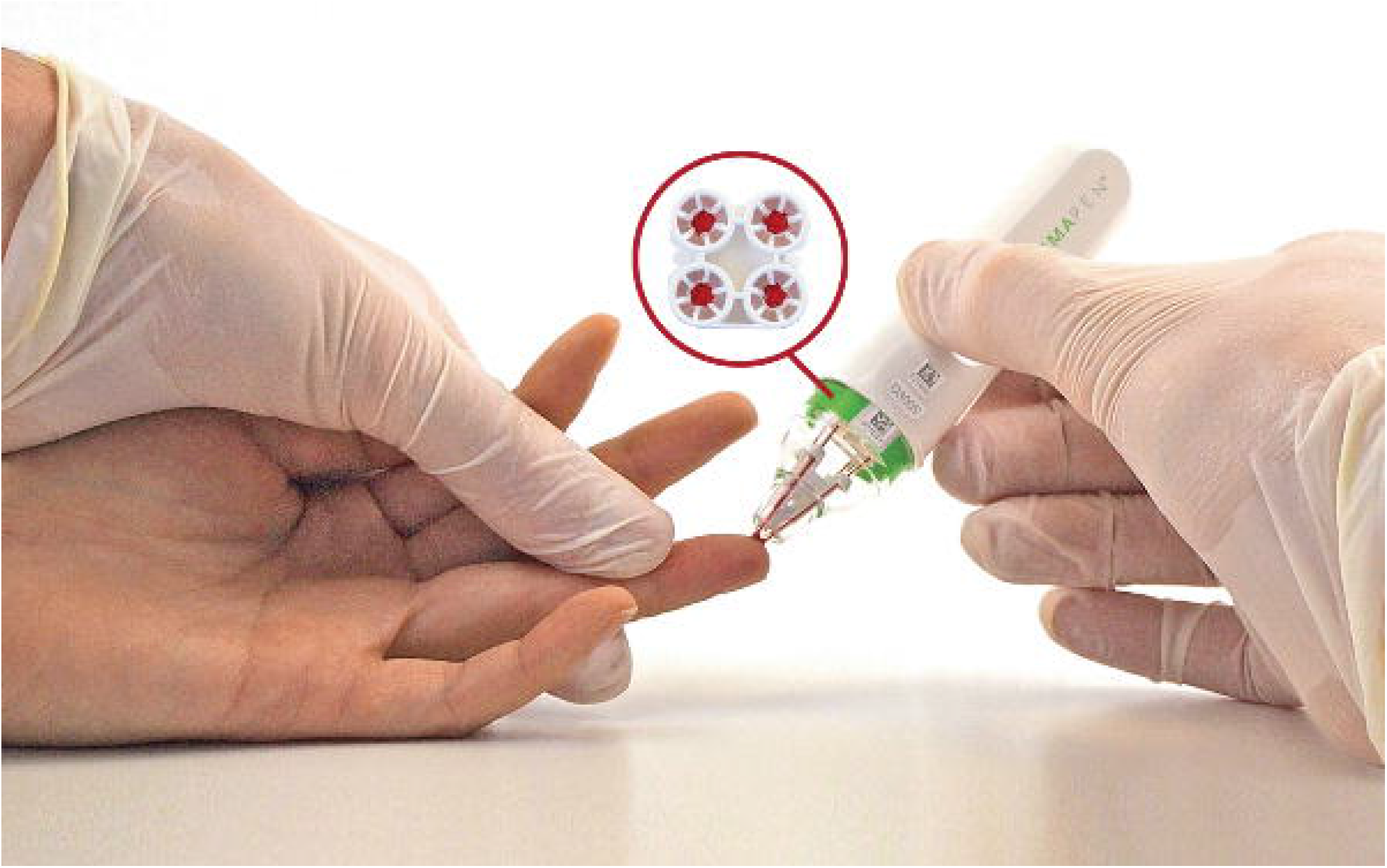
Illustration of the hemaPEN device in use. Following a fingerprick, blood is collected using the hemaPEN device onto four dried blood spot (DBS) discs. *Image acquired from Neoteryx by Trajan (https://www.neoteryx.com/media-download)*.

**Figure 2.**
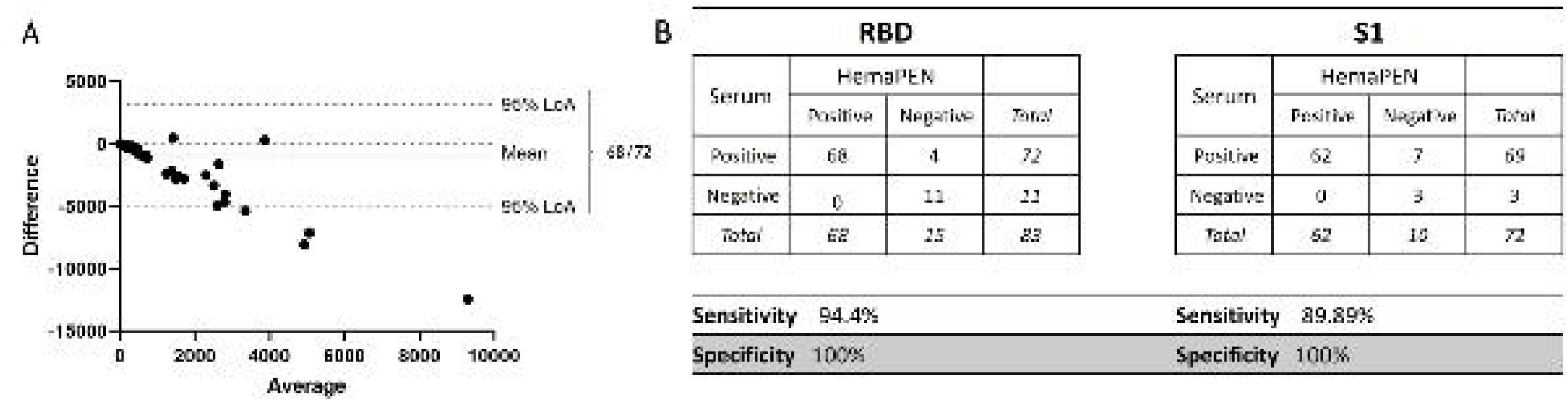
Comparison of paired hemaPEN DBS eluates and serum specimens for SARS-CoV-2 IgG specific for the RBD and S1 proteins of SARS-CoV-2. An in-house ELISA to measure IgG concentrations specific for the receptor-binding domain (RBD) of the SARS-CoV-2 spike protein was used to determine the number of seropositive samples (A), concentration of IgG (B), and a correlation (C) amongst paired hemaPEN DBS eluates and serum specimens. Seropositive results for RBD specific IgG were then assayed for IgG specific for the S1 antigen of the SARS-CoV-2 spike protein. Proportion of seropositive samples (D), concentration of IgG specific for S1 (E), and corresponding correlations (F) amongst paired specimens using a Pearson’s correlation test. A nonparametric Wilcoxon matched-pairs signed ranked test was used for statistical analysis with a two-sided P < .05 considered statistically significant. The cut-off for seropositivity is represented by the dotted line. OD: optical density. BAU: Binding antibody units. r: correlation coefficient.

### Comparison of SARS-CoV-2 antibody responses between hemaPEN DBS eluates and serum samples within children and adults

To assess whether there is a difference in the measurement of SARS-CoV-2 antibody response using the two sampling methods in paediatric and adult samples, we stratified our analysis into children and adult populations. Similar results were observed for both adults (Fig. 3A, C) and children (Fig 3B, D), with a significant decrease (P < .001) in SARS-CoV-2 RBD and S1 IgG response for hemaPEN DBS eluates compared to paired serum samples. COVID-19 vaccinated individuals had higher concentrations of RBD, and S1-specific IgG compared to unvaccinated group in both adult (Fig. 3A, C) and children (Fig 3B, D).

**Figure 3.**
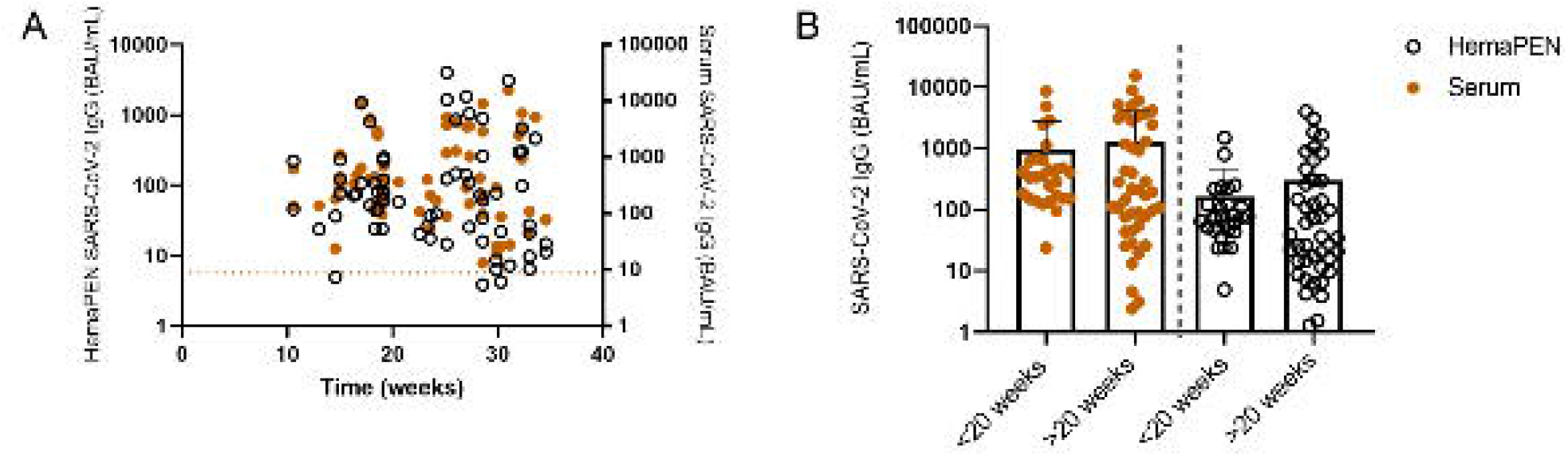
Comparison of SARS-CoV-2 IgG specific for the RBD and S1 protein in adults and children. An in-house ELISA was used to measure IgG concentrations specific for the receptor-binding domain (RBD) and S1 of the SARS-CoV-2 spike protein amongst paired hemaPEN DBS eluates and serum specimens in adults (A and C, respectively) and children (B and D, respectively). Individuals who have received at least one dose of COVID-19 vaccine was shown as gray lines, no COVID-19 vaccine shown as black lines and unvaccinated/infected with SARS-CoV-2 shown as blue lines. A nonparametric Wilcoxon matched-pairs signed ranked test was used for statistical analysis with a two-sided P < .05 considered statistically significant. The cut-off for seropositivity is represented by the dotted line. OD: optical density. BAU: Binding antibody units.

### Effect of storage time and temperature

To evaluate the stability of the DBS from the hemaPEN microsampling device, we compared IgG specific for S1 SARS-CoV-2 antigen between paired hemaPEN DBS eluates and serum samples over time (Fig. 4). Analysis of samples at MCRI occurred between 10-35 weeks (median 27 weeks) after sample collection (Fig. 4A). No significant differences in S1-specific IgG levels were observed when samples were assayed less than 20 weeks or more than 20 weeks after collection (Fig. 4B).

**Figure 4.**
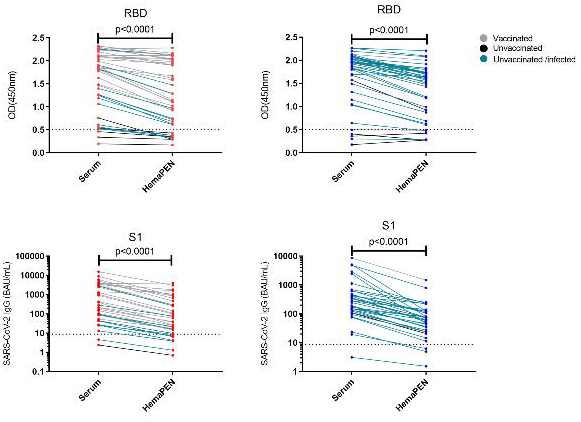
Effect of storage time on IgG specific for S1 protein for paired hemaPEN DBS eluates and serum samples. (A) hemaPEN DBS storage time and SARS-CoV-2 IgG concentrations in hemaPEN DBS eluates (open circle) or serum (orange circle) (B) SARS-CoV-2 IgG concentrations in hemaPEN DBS eluates (open circle) or serum (orange circle) stratified by storage time less than or greater than 20 weeks. Results are displayed as mean with standard deviation. A nonparametric Wilcoxon matched pairs signed ranked test was used for statistical analysis with a two-sided P < .05 considered statistically significant. The cut-off for seropositivity is represented by the dotted line. BAU: Binding antibody units.

### Sampling agreement between serum and hemaPEN DBS samples

A Bland-Altman analysis was used to compare the two blood collection methods where the ‘true’ value in the sample is unknown. Both sampling method displayed a high level of agreement with 95.18% of samples within the 95% limits (Fig. 5A). The sensitivity and specificity for the hemaPEN DBS eluates to measure SARS-CoV-2 RBD IgG was 94.40% and 100%, respectively when compared to serum specimens. For detection of S1 specific IgG, the hemaPEN DBS eluates had a sensitivity specificity of 89.89% and 100%, respectively.

**Figure 5.**
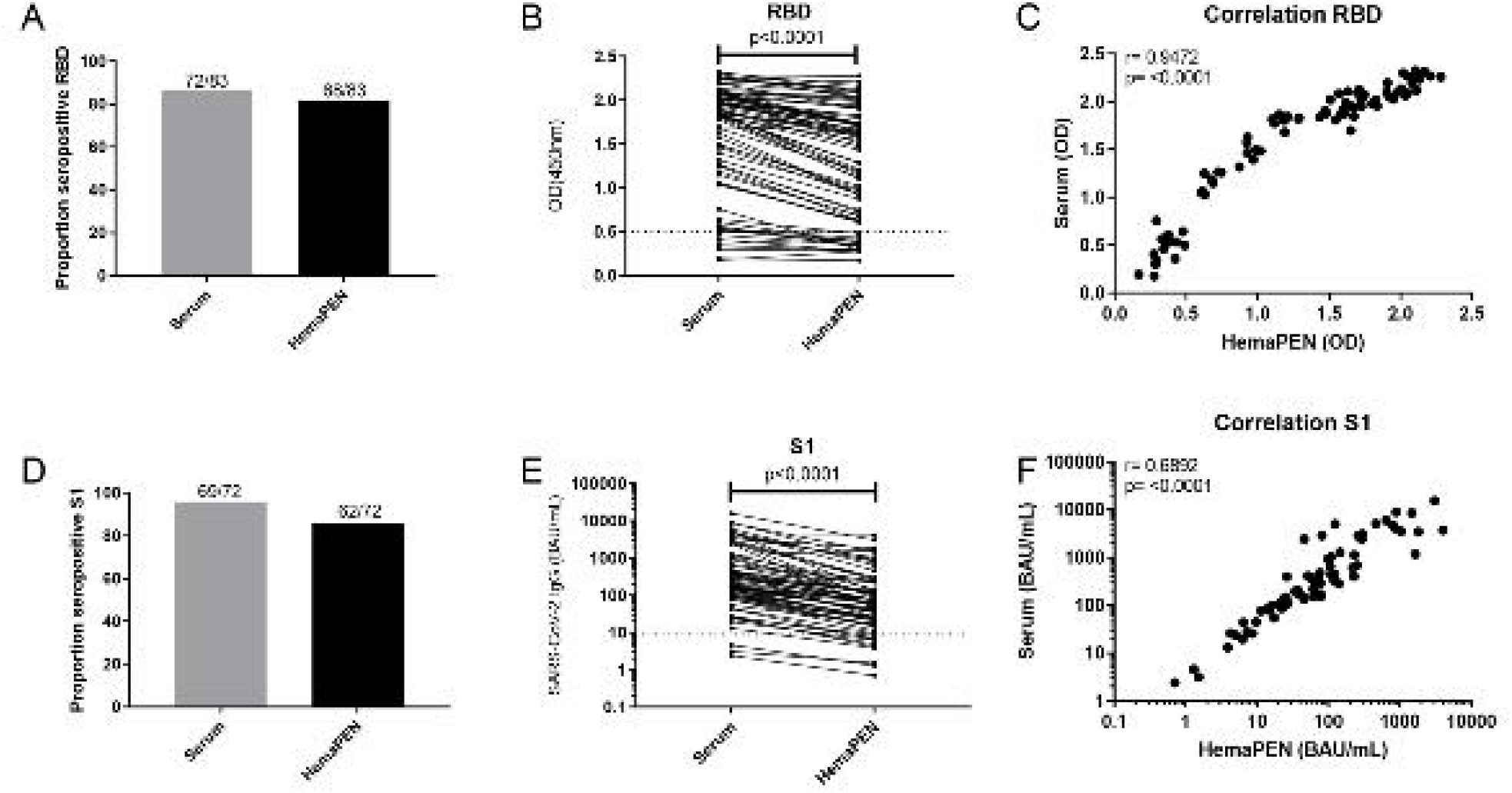
Bland-Altman plot of IgG concentrations specific for S1 antigen of SARS-CoV-2 (A) and sensitivity and specificity calculations amongst paired specimens for RBD and S1 specific IgG concentrations (B). The grey dotted lines represent the 95% limits of agreement.

### Synexa assay analysis on hemaPEN DBS specimens

Binding antibodies for the S1 SARS-CoV-2 protein was found to have a good correlation amongst the hemaPEN DBS eluates assayed on the Synexa assay versus paired hemaPEN DBS assayed at MCRI (Supplementary Fig. 1A, r = 0.6059, P <.0001). Seropositivity between binding S1 IgG assay platforms were comparable with no significant differences (Supplementary Fig. 1B). The Synexa also measured NAb based on competition with high OD representing a low NAb response. A negative correlation was observed with NAb and binding IgG antibodies with hemaPEN DBS eluates using the Synexa assay (Supplementary Fig. 1C, r = -0.7842, P <.0001). The proportion of positive NAb in the hemaPEN DBS eluates was higher in children (23/41; 56.1%) compared to adults (12/39; 30.8%) (Supplementary Fig. 1D).

## Discussion

Our results demonstrate that the DBS eluates from the novel sampling device, hemaPEN had homogeneity with paired venepuncture serum samples, and correlated well for detecting SARS-CoV-2 RBD, and S1 IgG. Despite the lower antibody concentrations observed in hemaPEN eluates when compared to serum, we found high sensitivity and specificity for the detection of SARS-CoV-2 spike antibodies in both paediatric and adult cohorts, providing confidence for the use of this microsampling device.

Our data indicates that hemaPEN sampling is best suited for individuals or populations with high levels of SARS-CoV-2 immunity, and may be used for serosurveillance to help inform public health responses. However, compared with serum samples, we found hemaPEN DBS eluates have lower sensitivity for samples with lower/borderline IgG concentrations that might result in false negative results. This was unexpected as we previously showed a strong correlation between SARS-CoV-2 RBD and S1 antibody levels between DBS collected via Guthrie card (Whatman 903 filter paper) and paired serum samples [8]. Other studies have also reported strong correlation of SARS-CoV-2 neutralising antibodies and binding antibody between DBS and paired serum samples [5, 13, 14]. However, comparison between studies are difficult as different elution buffers and elution volumes, as well as sample dilutions and type of antibody assays were used. For hemaPen DBS, further optimisation on the assay cut-offs and DBS elution volume may be needed to improve the sensitivity and specificity. In addition, it would also be interesting to examine whether DBS samples can be used to characterise antibody affinity and antibody-effector function.

Serosurveys remains an important component of the public health response against COVID-19 [15, 16]. Serosurveys monitor population immunity to SARS-CoV-2 and inform public health measures, including the administration of booster doses. The advantage of using DBS samples for serosurveillance is evident by providing a logistically more feasible approach, by reducing the need for a cold-chain transportation, highly trained personnel, and facilities. These factors are particularly relevant for remote settings in high-income countries and/or LMICs, or at-home settings for self-collection where there are logistical barriers. The hemaPEN device, offers an easier way to collect and store blood samples than traditional Guthrie card for immunological assessment. This is achieved through the capillary mechanism that enables the collection of volumetrically fixed samples, and for the samples to dry within the device reducing the risk of contamination [17-19]. Furthermore, we demonstrated acceptable stability of the hemaPEN DBS eluates to be stored for up to a minimum of 35 weeks at room temperature, a major advantage for remote settings. In terms of compatibility with existing laboratory infrastructure for processing of DBS specimens, no specialised equipment is needed for the hemaPEN device.

In addition to assessing sample sensitivity, evaluating the user experience and patient feasibility is important. Personal communication about the use of the hemaPEN device from both adult and parents of paediatric participants were well received due to its efficiency, and minimal blood collection, and was the preferred method of choice compared to venepuncture, particularly for children. Good clinical practice was instrumental in ensuring a successful and comfortable patient experience. Furthermore, the user-friendly nature of the hemaPEN microsampling device makes it a feasible option for self-collection applications when needed.

## Limitations

This study has a few limitations. The sample size was small, yet the advantage of this study was that it included both a paediatric and adult cohort. Furthermore, sample collection was undertaken opportunistically at either one or 12 months after the initial household member presented positive for COVID-19. This meant that samples were collected for antibody responses relative to time of COVID-19 infection, and not to time of COVID-19 vaccination. Nonetheless, the results observed in this study are consistent and demonstrates the validity of the hemaPEN device for the successful detection of SARS-CoV-2 antibodies.

## Conclusion

In conclusion, we showed that the hemaPEN device may be alternative to venepuncture for blood collection to measure SARS-CoV-2 immunity. This device will be relevant for serosurveillance studies and population (i.e., children) where a less invasive sampling is preferred. While we use this device to measure SARS-CoV-2 immunity, the device can also be used to monitor immunity to other infectious diseases, such as mumps and rubella, where DBS have been used, although testing against specific infectious agent is needed.

## Data Availability

All data produced in the present study are available upon reasonable request to the authors

## Acknowledgments

We thank the study participants and families for their involvement in this study. We would like to thank BioBank (MCRI) for processing the serum samples. We would also like to thank the researchers at Synexa Life Sciences for technical expertise. Funding for the recruitment of participants was provided by the Royal Children’s Hospital Foundation and the Infection and Immunity Theme, MCRI. PVL is supported by NHMRC Career Development Fellowship. This work is supported by Victorian Government’s Medical Research Operational Infrastructure Support Program. NWC received funding from the National Institute of Health for influenza and COVID-19 research and the Victorian Department of Jobs Precincts and Regions (DJPR).

## Author contributions

FL, NWC, ST and PVL were responsible for conceptualisation, investigation, project administration, visualisation, supervision, and funding. JR, BW, NJW, JD and AG were responsible for project administration, resources, investigation, and data curation. JN, RH, NM and ZQT were responsible for data curation, investigation, and methodology. NM and ZQT were also responsible formal analysis and writing-original draft. All authors were involved in writing-reviewing and editing the manuscript. The authors have accepted responsibility for the entire content of this manuscript and approved its submission.

## References

1. Koirala, A., et al., Paediatric SARS-CoV-2 serosurvey 2022, Australia Summary report 2022: The Paediatric Active Enhanced Disease Surveillance (PAEDS) Network and the National Centre for Immunisation Research and Surveillance (NCIRS).

2. Su, X., et al., Dried blood spots: An evaluation of utility in the field. J Infect Public Health, 2018. 11(3): p. 373–376.

3. Adam, O., et al., Seroprevalence of measles, mumps, and rubella and genetic characterization of mumps virus in Khartoum, Sudan. Int J Infect Dis, 2020. 91: p. 87–93.

4. Kaduskar, O., et al., Optimization and Stability Testing of Four Commercially Available Dried Blood Spot Devices for Estimating Measles and Rubella IgG Antibodies. mSphere, 2021. 6(4): p. e0049021.

5. Itell, H.L., et al., SARS-CoV-2 Antibody Binding and Neutralization in Dried Blood Spot Eluates and Paired Plasma. Microbiol Spectr, 2021. 9(2): p. e0129821.

6. Maritz, L., et al., Validation of high-throughput, semiquantitative solid-phase SARS coronavirus-2 serology assays in serum and dried blood spot matrices. Bioanalysis, 2021. 13(15): p. 1183–1193.

7. Meyers, E., et al., Diagnostic performance of the SARS-CoV-2 S1RBD IgG ELISA (ImmunoDiagnostics) for the quantitative detection of SARS-CoV-2 antibodies on dried blood spots. J Clin Virol, 2022. 155: p. 105270.

8. Toh, Z.Q., et al., The use of dried blood spots for the serological evaluation of SARS-CoV-2 antibodies. J Public Health (Oxf), 2022. 44(2): p. e260–e263.

9. Turgeon, C.T., et al., Detection of SARS-CoV-2 IgG antibodies in dried blood spots. Diagn Microbiol Infect Dis, 2021. 101(1): p. 115425.

10. Weisser, H., et al., Evaluation of dried blood spots as alternative sampling material for serological detection of anti-SARS-CoV-2 antibodies using established ELISAs. Clin Chem Lab Med, 2021. 59(5): p. 979–985.

11. Toh, Z.Q., et al., Comparison of Seroconversion in Children and Adults With Mild COVID-19. JAMA Netw Open, 2022. 5(3): p. e221313.

12. Tuckerman, J., et al., Seroprevalence of SARS-CoV-2 antibodies in health-care workers at a tertiary paediatric hospital. J Paediatr Child Health, 2021. 57(7): p. 1136–1139.

13. Miesse, P.K., B.B. Collier, and R.P. Grant, Monitoring of SARS-CoV-2 antibodies using dried blood spot for at-home collection. Sci Rep, 2022. 12(1): p. 5812.

14. Morley, G.L., et al., Sensitive Detection of SARS-CoV-2-Specific Antibodies in Dried Blood Spot Samples. Emerg Infect Dis, 2020. 26(12): p. 2970–2973.

15. Nair, D., et al., Sero-Surveillance to Monitor the Trend of SARS-CoV-2 Infection Transmission in India: Study Protocol for a Multi Site, Community Based Longitudinal Cohort Study. Front Public Health, 2022. 10: p. 810353.

16. Murhekar, M.V. and H. Clapham, COVID-19 serosurveys for public health decision making. Lancet Glob Health, 2021. 9(5): p. e559–e560.

17. Smidt, M., et al., Evaluation of hemaPEN(®) sampling device for measurement of cocaine and metabolites in capillary blood by LC-MS/MS. Bioanalysis, 2022. 14(20): p. 1295–1303.

18. Sen, A., et al., In vitro testing of the hemaPEN microsampling device for the quantification of acetaminophen in human blood. Bioanalysis, 2020. 12(24): p. 1725–1737.

19. Deprez, S., et al., Evaluation of the Performance and Hematocrit Independence of the HemaPEN as a Volumetric Dried Blood Spot Collection Device. Anal Chem, 2019. 91(22): p. 14467–14475.

