## Supplementary materials for "Evaluation of SARS-CoV-2 antibody response between paired fingerprick (hemaPEN®) and venepuncture collected samples in children and adults"

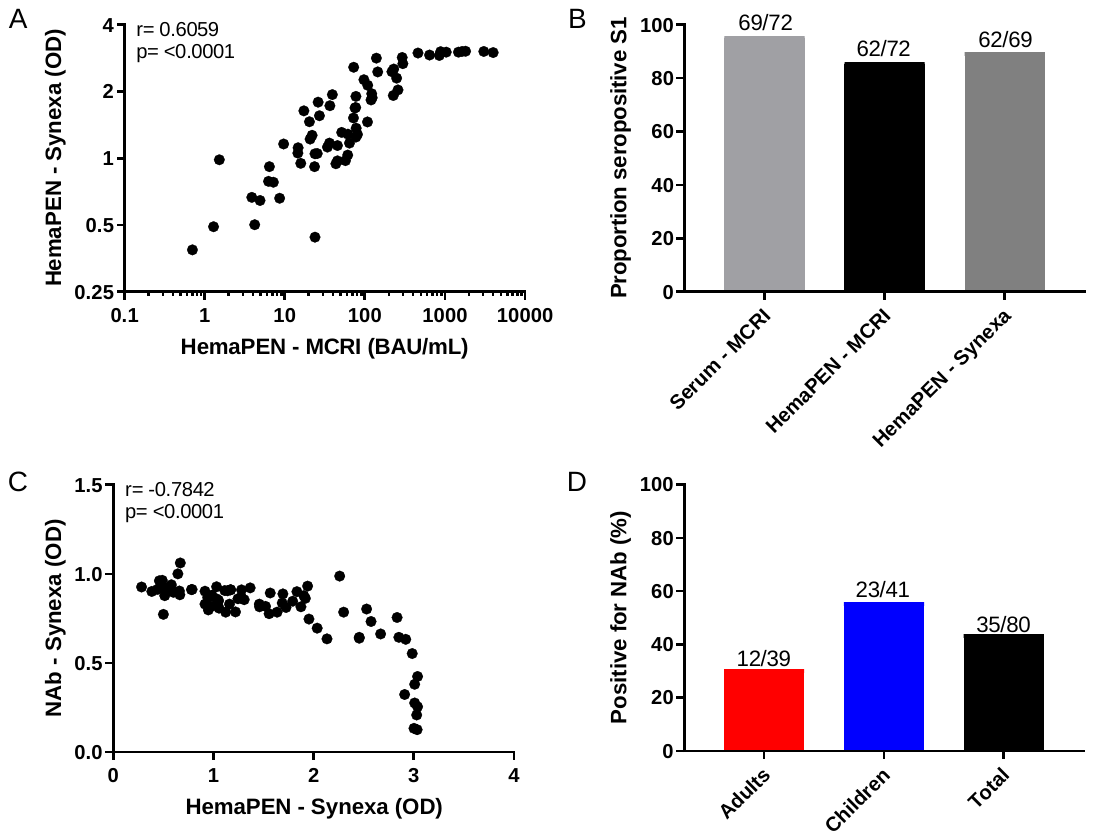

Supplementary figure 1. Synexa analysis of hemaPEN DBS eluates for binding IgG specific for S1 antigen of SARS-CoV-2 (A and B) and neutralising antibodies (NAb) (C and D). A Pearson’s correlation test was used for correlation analysis. OD: optical density. BAU: Binding antibody units. r: correlation coefficient.
